# How many COVID-19 PCR positive individuals do we expect to see on the Diamond Princess cruise ship?

**DOI:** 10.1101/2020.11.14.20230938

**Authors:** Jing Qin, Fang Chen, Huijuan Ma, Yukun Liu, Dean Follmann, Yong Zhou

## Abstract

The coronavirus disease 2019 (COVID-19) has become a global epidemic crisis with tens of thousands confirmed cases surfacing everyday. The infection rates in households, offices and public places are quite different from those in encompassed spaces such as airplanes, trains and cruise ships.

Studying the behavior of COVID-19 in confined spaces like Diamond Princess cruise is of great importance to understand the disease progression and to manage the epidemic. We propose a novel mixture model to estimate the infection distribution and total infected number after 14 days of quarantine based on PCR test data performed on the Diamond Princess cruise.

**Results:** In contrast to the officially reported 634 individuals with PCR-positive results after the 14 day quarantine, which as of April 27, 2020 had increased to 712, we conclude that this number should be at least 1000. The discrepancy might be caused by the false-negative result of the PCR test or the occurrence of infection after the test.

## Background

Since the outbreak of the novel coronavirus disease COVID-19, the epidemic has progressed rapidly in China and all over the world. As of June 2020, COVID-19 has emerged in more than two hundred countries and has caused an unprecedented global epidemic crisis with tens of thousands of new confirmed cases every day. One of the most well-known COVID-19 outbreaks is the eruption on the Diamond Princess cruise ship. The high contagiousness of COVID-19 on the Diamond Princess cruise has attracted global attention.

The Diamond Princess cruise ship started on January 20, 2020 in Yokohama, Japan and returned there on February 3, after visiting Kagoshima, Hong Kong, Vietnam, Taiwan, and Okinawa (Sekizuka *et al*., 2020). During this period, an 80-year-old passenger who disembarked on January 25 in Hong Kong, had presented with symptoms of a cough since January 23 and was confirmed positive for COVID-19 on February 1. The Japanese authorities doubted the widespread infection before the cruise ship went back, and asked the quarantine officers to investigate the health status of all passengers and crew members from on February 3-4 (Sekizuka *et al*., 2020). On February 5, there were 2,666 passengers and 1,045 crew members on board. To prevent further spread of the virus, the Japanese authorities asked 3,711 individuals, including passengers and crew members, to stay onboard in Yokohama to carry out a 14-day isolation period until February 19. During the 14-day quarantine, all passengers were required to stay in their own cabins, but crew members were not fully isolated in order to maintain limited services on the ship.

A confirmed case of COVID-19 infection was defined as a case with a positive result for testing in respiratory specimens. Quantitative reverse transcription polymerase chain reaction (qRT-PCR) or nested polymerase Chain Reaction (PCR) testing data (Sakiko *et al*., 2020) from February 5 to 20, including number of tests and number of testing positive cases each day, are publicly available. However, possibly due to the testing policy adjustment, the data on February 11 and 14 are not available. It is expected that the infection rates in the cruise were higher than those in any open places in other countries or districts. This might be caused by the confinement of the cruise, where individual isolation of all those aboard was not possible. Since the occurrence of the infection on Diamond Princess, at least 25 other vessels were also found high numbers of confirmed cases (Mallapaty *et al*., 2020). For example, in one Antarctic cruise, approximately 60% of passengers were confirmed cases (James *et al*., 2020). These unprecedented highly infectious events had harmful impacts on the cruise industry and resulted in an economic loss of more than one billion US dollars. Moreover, on April 18, 2020, French Defense Minister Florence told lawmakers that more than 1,000 of the 2,300 people aboard the Charles de Gaulle aircraft carrier had tested positive (Jeffrey *et al*., 2020). COVID-19 cases were also observed on multiple aircraft carriers in USA. Therefore, the research on the transmissibility pattern of COVID-19 on highly populated confined spaces like the Diamond Princess is indispensable.

The collected data on the Diamond Princess have unique features because at the beginning, the upper-respiratory specimens were collected from symptomatic passengers, crew, and their close contacts for PCR testing. Starting February 11, due to the expansion of laboratory capacity, quarantine officers systematically collected respiratory specimens from all passengers by age group, starting with those aged 80 years and older as well as individuals with comorbidities, such as diabetes or a heart condition. This means the PCR testing was conducted according to individual characteristics and not at random. Therefore, it is desirable to use different techniques to model the two-part data separately. It is critical to know how the infection varies from quarantine onset in order to avoid being infected. A sensible estimate of infection distribution plays a vital role in epidemiology studies. Our goal is to model the infection patterns of Diamond Princess such as estimating the number of infected individuals when released on February 20 and the infection onset distribution on the Diamond Princess cruise ship. To the best of our knowledge, current global research on cruise ships have scant statistical analyses on transmission of COVID-19, thus ongoing efforts to model epidemic data from the Diamond Princess will provide important information on how the spread of COVID-19 in a cruise ship differs from that in open spaces. Understanding the extent of transmission of novel coronavirus in a closed environment has major implications for controlling and anticipating the trajectory and impact of this pandemic, and for the prevention and control of similar diseases in general. A better understanding of the virus transmission pattern might also give some insights on how to prevent the transmission of virus, making the cruise travel safer in the future.

Daily time series of RT-PCR test data from February 5 to February 20 can be found in Table 1 of Mizumoto et al.(2020), including the number of tests, number of testing positive cases, number of cases in presence or absence of symptoms, etc (Mizumoto *et al*., 2020). However, the infection time and the infection rate were unknown. Data from February 11 and February 14 were not available in the original data sources. We carefully address this missing data problem in our statistical analysis. At the beginning, PCR tests had been conducted mainly for symptomatic groups and their high-risk close contacts, and then for almost all persons in the second week. As of February 20, a total 3063 respiratory specimens were tested with 634 positive, including one quarantine officer, one nurse, and one administrative officer. Of these 634 cases, 313 were female and 321 were male. 476 cases had age 60 years or older (Mizumoto *et al*., 2020). For convenience, the daily time series data with number of tested individuals and individuals with positive results are provided in Table 1.

**Table 1:**
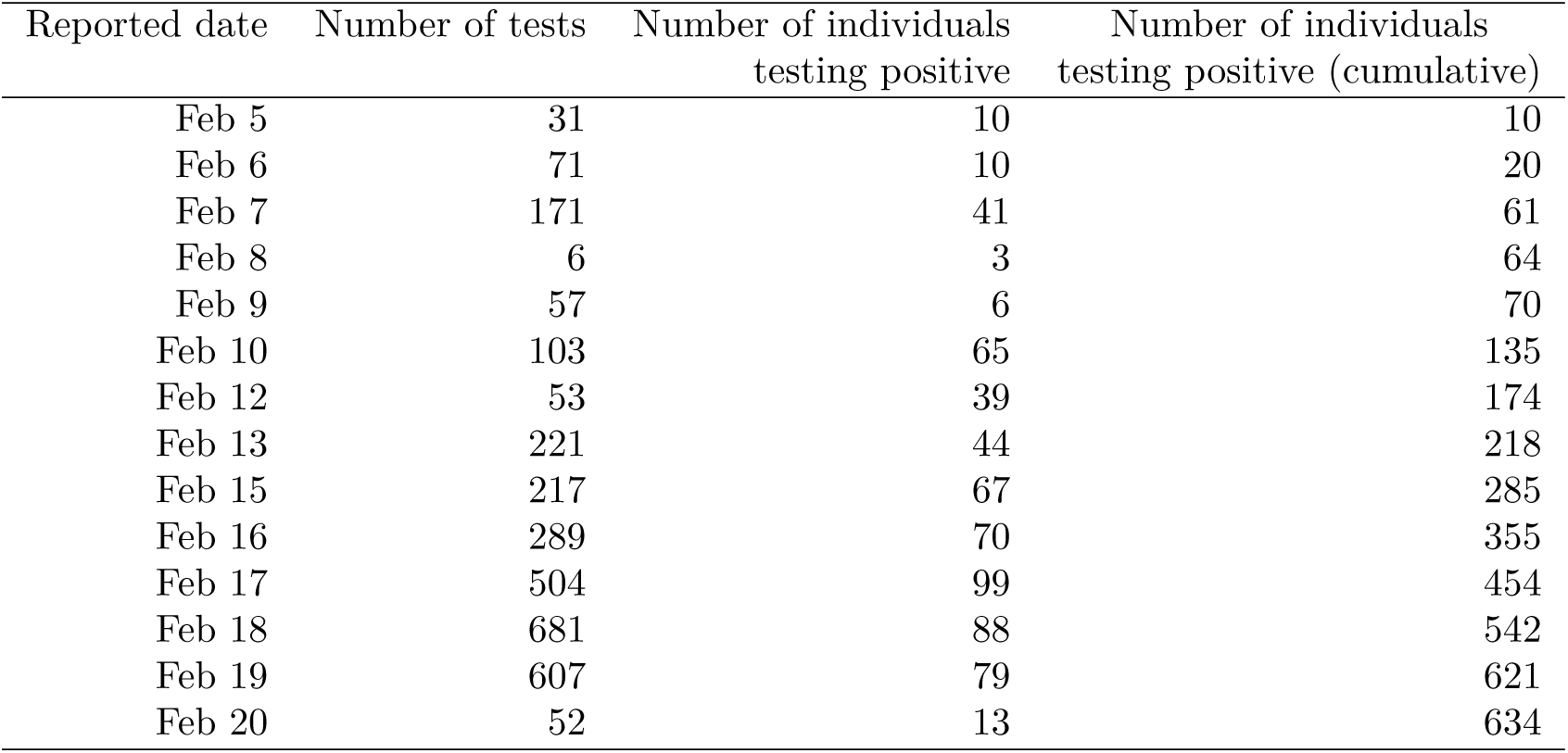
Number of tests and number of individuals testing positive for passengers and crews on the Diamond Princess cruise ship, Yokohama, Japan, February 2020 (n = 3711). Data on February 11 and 14 are not available.

In this article, we propose a cure mixture model to characterize the epidemic infectious pattern of Diamond Princess ship, which combines Weibull distribution assumption for the infection time with logistic form of the selective probability. Based on the maximum likelihood approach, the infection distribution and the estimated total number of infections on Diamond Princess at the end of 14 days quarantine are given. We conclude that undocumented infections might exist based on our analysis.

## Statistical methods

In this Section, we provide an obvious lower bound estimation of the number of infected individuals and a more sophisticated statistical method to estimate this number and the infection distribution using the daily time-series data described in Table 1. Let *f*(*x*) and *F*(*x*) be the density and cumulative distribution functions of infection onset time *X* calculated from February 4, respectively. Therefore *F*_*i*_ = *F*(*i*) is the probability of the infection onset time occurring before day *i* starting from February 4. For example, *F*_1_ represents the probability of testing positive on February 5. According to the non-decreasing property of the distribution function, *F*_*i*_ should satisfy the constraint *F*_1_ ≤ *F*_2_ ≤ … ≤ *F*_14_.

First, we may hypothetically assume that no PCR tests were performed in the first 13 days of quarantine. On February 20, the last quarantine day, 52 individuals were selected by the officer on the ship for testing and 13 positives were found. Therefore the infection rate is 13*/*52 = 25%. Out of the *n* = 3711 passengers and crew members, we expect to see 3711 *\** 0.25 = 928 PCR positive results. In reality, the selection of individuals for PCR test in the first week was not random. Symptomatic individuals, elders and closely related individuals were selected first. Therefore, 928 should be a lower bound estimation of total PCR-positive individuals after the end of quarantine, as these 52 should be less likely to be PCR-positive compared to the ship as a whole. We will give a full explanation of the larger estimation of N theoretically below.

Next we shall show statistically that a non-random sampling was implemented in the selection for PCR test. Suppose there is no selection bias, i.e., a random sampling was used. Let Let *n*_*i*_ and *N*_*i*_ be the number of tested positive cases and number of tests at day i respectively, and *X*_*ij*_ be the infection time of the *j*th subject who was tested at day *i, j* = 1, 2, …, *N*_*i*_; *i* = 1, 2, …, 14. Instead of observing the exact infection time *X*_*ij*_, we can only observe the number of tested positive individuals *n*_*i*_, which is equal to 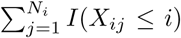 with conditional expectation *E*(*n*_*i*_) = *N*_*i*_*F*_*i*_, Let *δ*_*ij*_ = *I*(*X*_*ij*_ ≤ *i*), then 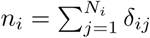. We use the nonparametric likelihood method directly to estimate *F*_*i*_. The observed likelihood function is

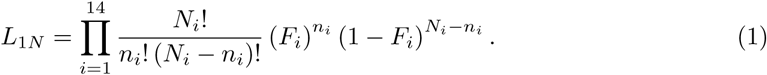

This is a standard current status data problem discussed extensively in statistical literature, for example, Sun (2006). To maximize the log-likelihood

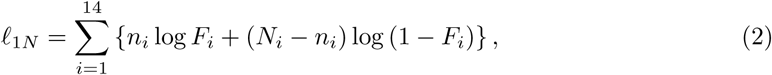

with non-decreasing constraints on *F*_1_ ≤ *F*_2_ ≤ … ≤ *F*_14_, we can solve the above constrained optimization problem using the well-known pool adjacent violators algorithm (PAVA) invented by Ayer et al.(1955), which has been implemented in R packages like Iso or isotone. Easily we can find again 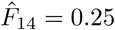. Therefore the estimation of the number of PCR-positive patients remains the same. However, frequency and estimated probability on each day have a large discrepancy, especially in the first week. This demonstrates that the random selection process was violated.

Next we provide a novel mixture modeling strategy that fully utilizes the non-random sampling method adapted in the Diamond Princess cruise during the 14-day quarantine. Although COVID-19 is a very contagious disease, some people may be immune to it. Naturally the cure rate model discussed by Farewell (1982), for example, can be used.

At each day *i*, we suppose that tested individuals are a mixture of a proportion of *λ*_*i*_ susceptible individuals who eventually get infected and a proportion of 1 *− λ*_*i*_ who are not susceptible to COVID-19 and never get infected on the cruise. This means,

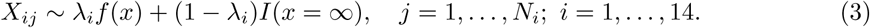

Motivated by the priority in choosing symptomatic or high-risk groups, the suspectable probabilities in the first week maintained a high level defined as *λ*, which is estiamted approximately equal to 1 by MLE method. Since symptomatic and vulnerable individuals were tested first and some sick individuals disembarked at the end of first week, it is expected that the proportions of non-susceptible individuals became larger as time went by. In other words, starting from the second week, *λ*_*i*_, *i* = 8, 9, …, 14 contain a downward trend. We suppose the mixture proportion *λ*_*i*_ varies across *i* in the logistic form to add model flexibility. And we assume a Weibull distribution for *F* which is commonly used in epidemical modelling. Specifically, suppose

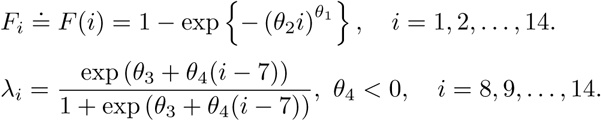

where *θ*_1_, *θ*_2_ are separately the shape, scale parameters of weibull distribution and coefficients *θ*_3_, *θ*_4_ characterize the logistic form. It is easy to see that the suspectable probabilities *λ*_*i*_, 8 ≤ *i* ≤ 14 in the second week have a logistic regression form and decrease as *i* increases. This means, we assume the proportion of unsusceptible individuals in the second week increases, i.e., more and more unsusceptible subjects were left to be tested.

At this time, we have *E*(*n*_*i*_) = *λ*_*i*_*N*_*i*_*F*_*i*_. The observed likelihood function is

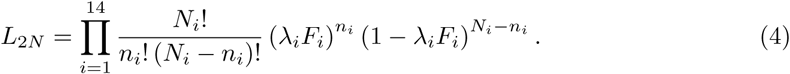

We propose to maximize the following weighted log-likelihood

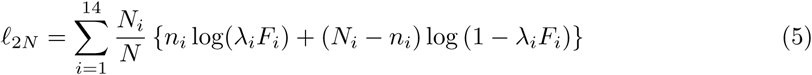

with respect to the underlying parameters, where 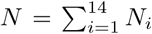 is the total number of tested individuals during the quarantine period. The maximum likelihood estimators (MLEs) 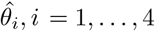 can be obtained by

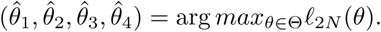

Let *n* = 3711 be the total number of passengers and crews onboard on February 5. Denote

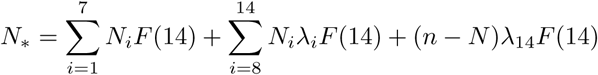

as the theoretical number of PCR positive individuals at the end of 14 days quarantine, which can be estimated by

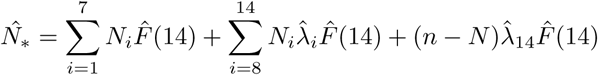

In the above formulaiton, 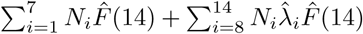 represents the estimated number of PCR positive cases among individuals tested before February 20 and 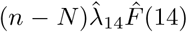 is the predictive infectious value for those conducted PCR tests later.

## Application to Diamond Princess Cruise Ship

We apply the proposed parametric mixture model to PCR test data performed on the Diamond Princess and maximum likelihood estimators of parameters are obtained. We used a parametric bootstrap approach to derive confidence intervals of the unknown parameters. Specifically, we generated 500 bootstrap samples based on the parametric mixture model with estimated parameters. The 95% confidence intervals (CI) were derived through normal approximation, where the estimated standard errors were calculated as the standard deviation of bootstrap sample estimators.

Using the mixture modelling strategy, the maximum likelihood estimators by maximizing the joint log-likelihood with time series daily report data are 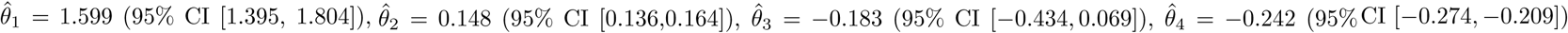. The density function *f*(*x*) and cumulative distribution function *F*_*i*_ can be estimated by the Weibull distribution with estimated parameters. *λ*_*i*_*F*_*i*_ can be obtained along the same line, where 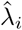 is the estimated proportion of susceptible individuals incorporated in the PCR test at day *i*.

Based on our novel mixture model, the estimated total infected number *N* at the end of quarantine is 1036, with 95% confidence interval [970, 1103]. We estimated *N* by combining the total tested number 3063 before February 20 with 3711*−*3063=648 individuals whose test results were unknown. The estimated number 1036, is larger than the obvious lower bound estimation 928 in the previous section, which is in accordance with our expectation. Figure 2 displays the estimated Weibull density function that specifically shows the infection pattern among susceptible individuals on the cruise. It is easy to see from Figure 2 that the infection rate among susceptible individuals rises at the first three to four days, and reaches the maximum at February 8, then decreases gradually. The infection rates in the first week were much higher than those in the second week.

**Figure 1:**
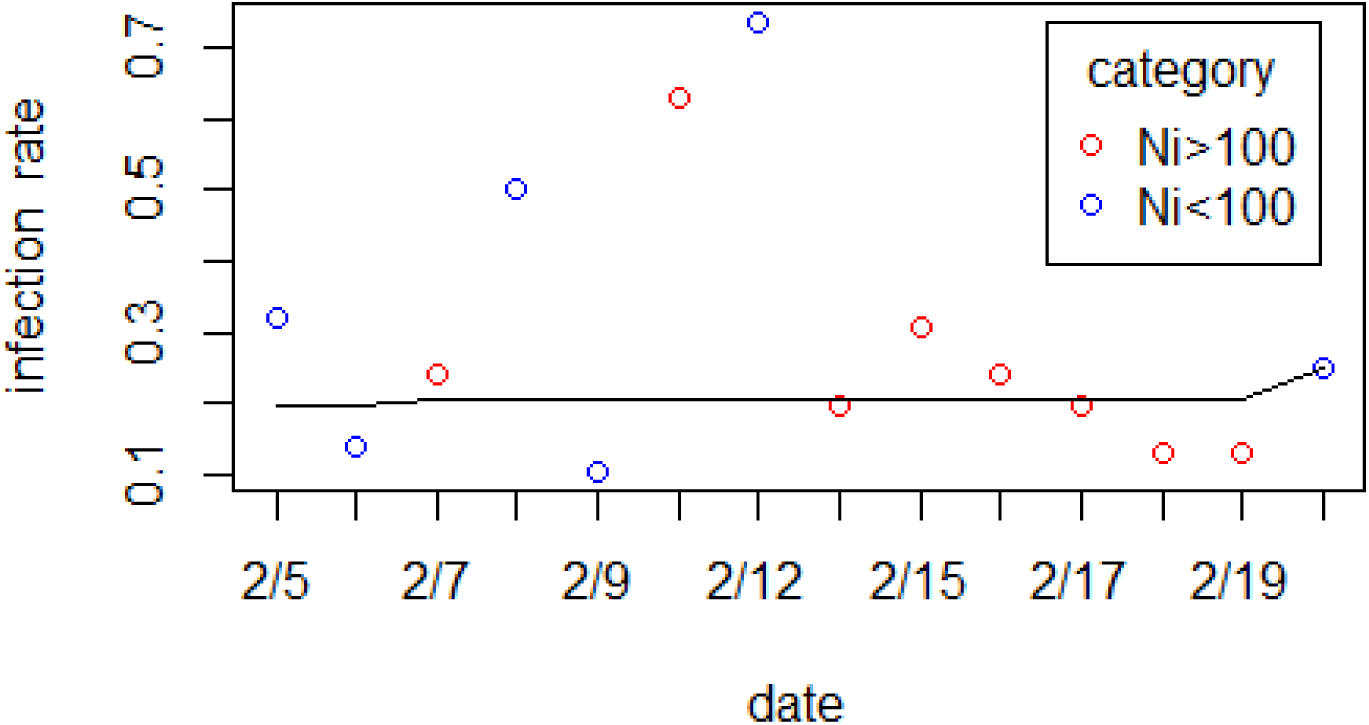
Comparison of **n_i_*/*N_i_** and fitted infection rates using PAVA. *n*_*i*_ represents daily number of positive individuals, *N*_*i*_ is the total number of tests each day. Scatter points are the rate of *n*_*i*_*/N*_*i*_. Blue points represent those whose corresponding total test number was less than 100, while red points mean the total test number was larger than 100. Black line shows the fitted infection distribution consisting of *F*_*i*_.

**Figure 2:**
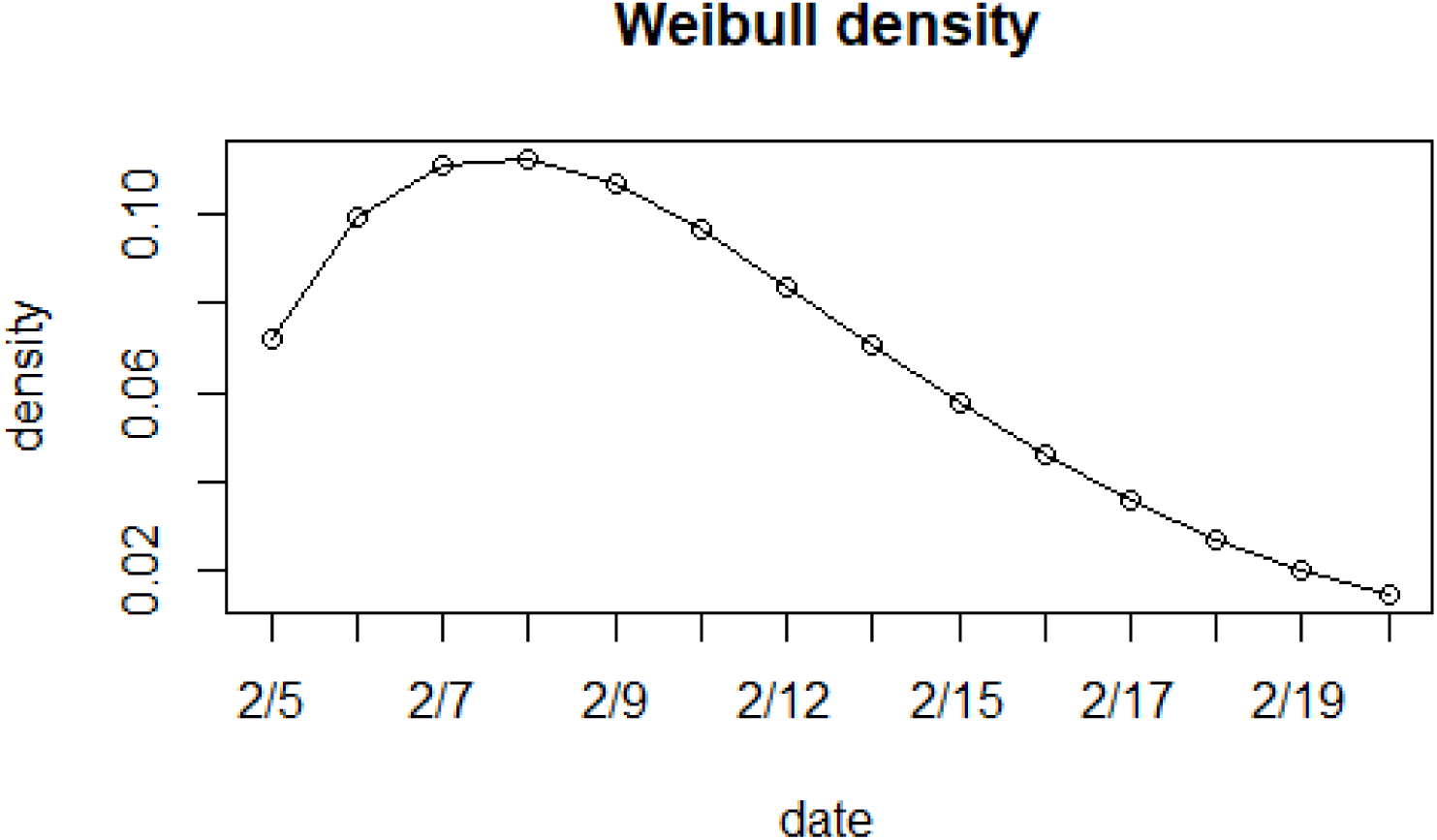
Fitted Weibull density function of infection time

Figure 3 visualizes the fitted infection proportions (black solid line) along with the scatter points of daily observed infection proportions *n*_*i*_*/N*_*i*_. We can see from Figure 3 that the estimated positive ratios fit the observed positive-test rates *n*_*i*_*/N*_*i*_ quite well, because the daily estimates of infection proportion increased rapidly in the first week and went down day by day in the second week. We obtained the mean infection time among susceptible individuals was at day 6.06 of the quarantine. Moreover, in this scatter plot, we used red and blue to demonstrate the total number of tests *N*_*i*_ *>* 100 or not, respectively. We found the estimated infection proportions were a better fit for *N*_*i*_ *>* 100. This is not surprising because more weights should be given to points where *N*_*i*_ *>* 100 with larger numbers of tested individuals. Points where *N*_*i*_ *<* 100 with small tested individuals might cause statistical instability. It is worth pointing out that only 6 people were tested on February 8 with 3 positive tests, which made the infection rate observed on that day to be as high as 50%. The small sample problem causes the instability in statistical analysis.

**Figure 3:**
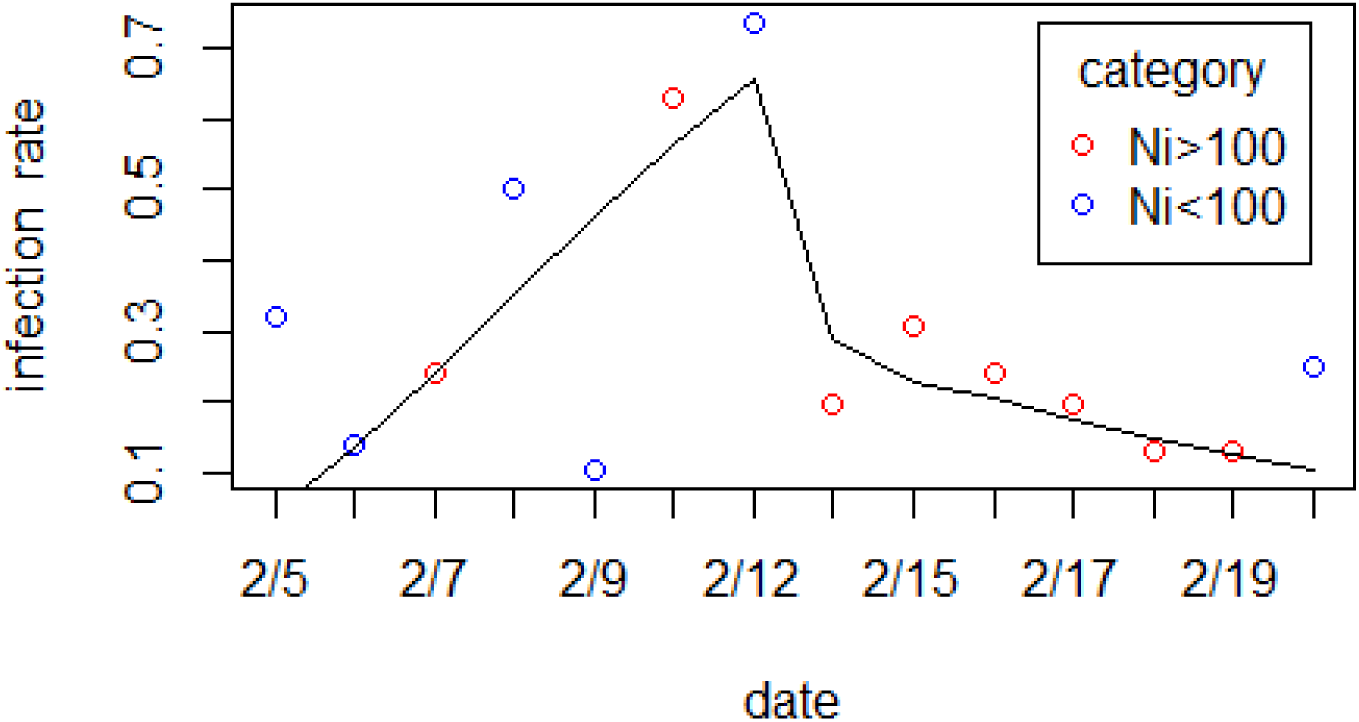
Comparison of n_i_*/*N_i_ and fitted weibull infection distribution. *n*_*i*_ represents daily number of positive individuals, and *N*_*i*_ is the total number of tests each day. Scatter points are the rate of *n*_*i*_*/N*_*i*_. Black line shows the fitted infection distribution consisting of *λ*_*i*_*F*_*i*_.

## Discussion

According to the public available data released by Johns Hopkins University, there were 712 confirmed cases on Diamond Princess cruise ship as of April 6, 2020. Our “obvious” estimate of 928 by using the last day quarantine data and mixture modelling based estimation of 1036 by taking the biased selection process into account both suggest that the number of PCR-positive individuals should be larger than the reported one. Below is some evidence that may potentially support our findings of the larger estimated *N*.

Hung (2020) recruited 215 passengers from Hong Kong who had been on board the Diamond Princess cruise ship. All 215 participants had been found to be negative for SARS CoV-2 by PCR 4 days before disembarking and were transferred to further quarantine in a public estate in Hong Kong, where they were recruited. Participants were prospectively screened by quantitative PCR and other detection methods during the quarantine. Of these 215 participants, nine individuals were positive for SARS-CoV-2 by RT-qPCR or serology while they were considered to be “uninfected” on the cruise. The unprecedented spread of COVID-19 owes its high transmissibility of pre-symptomatic and asymptomatic transmission. A large fraction of infections is asymptomatic and many others result in mild symptoms that could be mistaken for other respiratory illnesses (Perkins *et al*., 2020). Especially at the beginning of global spread in February when epidemiological recognition of viruses was insufficient, accuracy of medical tests for COVID-19 were limited. There are several infected individuals who had tested negative several times before finally being confirmed positive. Reported by Daily Shincho, a doctor treated coronavirus infections at the National Center for Global Health and Medicine of Japan claimed that the sensitivity of PCR test was about 70%. Sensitivity is an index to measure the proportion of positives, in terms of 70%, 30% of actual positives may be judged as false negatives. In fact, there were a series of people in Japan and abroad who tested positive after disembarking, even though they tested negative on board. Any passengers and crew members might test negative at their first examination and stayed on board until quarantine ended. The confined virus transmission environment on a cruise provides high risk of infection within these people, particularly for asymptomatic individuals who passed unnoticed even when they disembarked at the end of quarantine.

Using data before February 16, a recent paper by Zhang et al.(2020) used their estimated reproductive number *R*_0_ to estimate total number of 1514 confirmed cases. This, together with our findings, indicates that other transmission routes may be neglected, such as aerosol transmission via the central air conditioning system or drainage systems (Zhang *et al*., 2020). These factors point to a potentially large reservoir of unobserved infections.

Our statistical methods focus on exploring the information contained in the data, without regard to the impact of unnoticed incidents. For example, it should be noted that some asymptomatic cases may be missed due to the imperfect sensitivity of the PCR test. As pointed by Zhao et al.(2020), PCR-based viral RNA detection is currently the main way to diagnose COVID-19 infections in practice. Many cases strongly epidemiologically linked to novel coronavirus exposure with typical lung radiological findings remained RNA negative in their upper respiratory tract samples (Zhao *et al*., 2020). In addition, Arevalo-Rodriguez et al.(2020) suggest that repeated testing in patients with suspicion of COVID-19 infection is necessary given that up to 29% of patients could have an initial RT-PCR false-negative result. False negative rate of PCR based SARS-CoV-2 tests has been studied, recently, by Zhao et al.(2020), Liu et al.(2020) and Kucirka et al.(2020). We recommend that serological testing results on the Diamond Princess members can be included to make sure how many subclinical infections there really were.

The generalizability of transmission in confined circumstances of the cruise revealed how COVID-19 could spread in the community. Data from the early phase of local outbreaks are particularly powerful for predicting the impact of this virus. Based on our statistical analysis, we conclude that novel coronavirus spread in the cruise ship is easier and faster than that in open spaces. This result, together with extensive pre-symptomatic and asymptomatic transmission of COVID-19, suggests that a large number of unobserved infections might exist. Mizumoto et al.(2020) have estimated that 17.9% of the confirmed cases on Diamond Princess cruise were asymptomatic. After closure of the Huanan Seafood Wholesale Market on January 1 2020, within 240 cases with no exposure to it, 200 individuals (83%) reported had no direct contact with any individuals with respiratory infections (Ferretti *et al*., 2020), implying that many individuals were infected by pre-symptomatic or asymptomatic individuals. To analyse 40 primary-secondary pairs both with known onset of symptoms, Ferretti et al.(2020) found that nearly 37% of them belong to pre-symptomatic transmission. Direct transmission through a contact might occur at a time before awareness of taking conservatory measures. Meanwhile, exposure of the infected mainly depends on symptom-based surveillance all over the world. Many individuals linked to an identified source can be discovered by retrospective analysis. Tracing back to presumed exposure or a location where virus spread is a good approach to explore more unnoticed infections.

## Conclusion

In this paper we have explored two statistical methods and mainly focused on a novel mixture model to quantify the infection distribution and the susceptible proportions among daily tested people. Since the daily numbers of infected cases are not observable, it is worth pointing out that our statistic modelling and inference play an important role to estimate them accurately. Based on these estimates, the number of estimated cumulative infections at the end of quarantine is larger than the reported number 712. The novel coronavirus spread in the cruise ship is easier and faster than that in open spaces. The confined circumstances on the cruise are more beneficial to the transmission of virus. The high contagiousness of undocumented human infections also can result in both nosocomial and community transmission. Thus under the overall control of the current epidemic situation, long-term and continuous preventive measures are paramount to avoide the second outbreak of the epidemic.

## Data Availability

Data are available.

## Acknowledgements

We thank Benjamin Snow, ELS, from Leidos Biomedical Research, Inc for providing a technical review of the manuscript.

## Notes

### Competing Interest Statement

The authors have declared no competing interest.

### Funding Statement

This research is supported by the National Natural Science Foundation of China (11901200, 71931004, 11771144), Shanghai Pujiang Program (19PJ1403400)

### Author Declarations

East China Normal University, China

